# Repetitive but not single blast mild traumatic brain injury increases ethanol responsivity in mice and risky drinking behavior in combat Veterans

**DOI:** 10.1101/2020.11.10.20229427

**Authors:** Abigail G. Schindler, Britahny Baskin, Barbara Juarez, Suhjung Janet Lee, Rebecca Hendrickson, Katherine Pagulayan, Larry S. Zweifel, Murray A. Raskind, Paul E.M. Phillips, Elaine R. Peskind, David G. Cook

## Abstract

Mild traumatic brain injury (mTBI) is common in civilians and highly prevalent among military Servicemembers and in contact sports athletes. mTBI, especially within military populations, is often comorbid with posttraumatic stress disorder (PTSD), and can increase health-risk behaviors (e.g., sensation/novelty seeking, impulsivity, risk taking, irritability/aggression) and substance misuse/abuse, but underlying mechanisms remain poorly understood. Using an established mouse model of blast mTBI, here we examined the effects of single (1x) and repetitive (3x) blast exposure on ethanol responsivity using a battery of tests, each associated with distinct aspects of alcohol abuse vulnerability. While both single and repetitive blast exposure increased the sedative properties of high-dose ethanol (with no change in tolerance or metabolism), only repetitive blast exposure potentiated ethanol-induced locomotor stimulation and shifted ethanol intake patterns (i.e., increased consumption ‘front-loading’) during intermittent two bottle choice. To establish translational relevance, we next examined self-report responses to the Alcohol Use Disorders Identification Test-Consumption Questions (AUDIT-C) and used a novel unsupervised machine learning approach to investigate whether a history of blast with acute symptoms and mTBI affected drinking behaviors in Iraq and Afghanistan Veterans. AUDIT-C scores were increased in Veterans with a history of blast exposure and subsequent cluster analysis identified a three-cluster solution: ‘low’ (low intake and low frequency), ‘frequent’ (low intake but high frequency), and ‘risky’ (high intake and high frequency), where Veterans with a history of blast mTBI displayed a shift in cluster assignment from ‘frequent’ to ‘risky’, as compared to Veterans who were deployed to Iraq and/or Afghanistan who had no lifetime history of TBI. Together, these results offer new insight regarding how blast mTBI may give rise to increased substance use/misuse and highlight the increased potential for adverse health-risk behaviors following repetitive blast mTBI exposure.

## INTRODUCTION

Traumatic brain injury (TBI) affects every segment of the population and can be comorbid with post-traumatic stress disorder (PTSD), leading to significantly decreased quality of life and social and occupational functioning. This is particularly important for Veterans, who experience disproportionate rates of physical and mental illness compared to civilian counterparts [1-3]. Critically, US Veterans of the conflicts in Iraq and Afghanistan (Operation Enduring Freedom/Operation Iraqi Freedom/Operation New Dawn (OEF/OIF/OND)) exhibit especially high rates of mild traumatic brain injury (mTBI; labeled the “signature injury” of these conflicts) that is highly comorbid with PTSD [4-7]. OEF/OIF/OND mTBI rates are estimated at 10-25% (approximately 400,000 Veterans diagnosed since 2000), with PTSD comorbidity estimated at 50-75% [4-7]. Most commonly repetitive in nature, blast exposure (via detonation of high explosives) is the primary source of mTBI in this population, a significant driver of PTSD, and a major source of morbidity among Veterans enrolled in the VA health care [5, 7-9]. Both mTBI and PTSD are associated with increased health-risk behaviors (e.g., sensation/novelty seeking, impulsivity, risk taking, irritability/aggression) [5, 10-21] and substance use/misuse (e.g., alcohol, marijuana) [19, 20, 22, 23], potentially compounding negative outcomes following injury/trauma, but underlying mechanisms remain poorly understood.

Previous work in animal models supports the notion of brain injury as a risk factor for adverse health-risk behaviors, including substance misuse/abuse [18, 24-32]. Indeed, using an established mouse model of blast exposure [18, 32-36], we previously demonstrated increased novelty-seeking behavior and potentiation of stimulated phasic dopamine release within the nucleus accumbens [18]. These results highlight a potential underlying mechanism for the increase in health-risk behaviors seen following mTBI, as this brain region and neuromodulatory system are highly implicated in the motivating and rewarding properties of abused substances. Conversely, results from other mTBI models (i.e., impact, blast with body shielding) reported increased ethanol-induced sedation and decreased voluntary ethanol consumption [25, 27], highlighting the possibility that increased ethanol sedation functions to limit voluntary intake levels. Critically, no study to date has examined whether blast or impact mTBI affects ethanol-induced locomotor stimulation (a behavioral paradigm with implications for both appetitive and aversive ethanol actions) and/or whether repetitive trauma exposure produces disparate outcomes from only a single mTBI.

Here we investigated in mice potential effects of single (1x) and repetitive (3x) blast exposure on low-dose ethanol-induced locomotor stimulation, high-dose ethanol sedation, tolerance, and metabolism, and finally, voluntary ethanol intake (intermittent two bottle choice). Using an electronically-controlled pneumatic shock tube that models battlefield-relevant open-field blast forces generated by detonation of high explosives [18, 32-36], we found that while both single and repetitive blast exposure increased the sedative properties of high-dose ethanol (with no change in tolerance nor metabolism), only repetitive blast exposure resulted in potentiation of ethanol-induced locomotor stimulation and a shift in ethanol intake patterns (i.e., increased consumption ‘front-loading’) during intermittent 2BC. In order to establish translational relevance, we next examined self-report responses to the Alcohol Use Disorders Identification Test-Consumption Questions (AUDIT-C) [37] in OEF/OIF/OND Veterans without alcohol use disorder and used a novel unsupervised machine learning approach to investigate whether a history of repetitive blast exposure and mTBI affected drinking behaviors. Indeed, we found increased AUDIT-C scores in OEF/OIF/OND Veterans with a history of blast with acute symptoms, supporting the translational relevance of our animal model results. Subsequent AUDIT-C cluster analysis identified a three cluster solution: ‘low’ (low intake and low frequency), ‘frequent’ (low intake but high frequency), and ‘risky’ (high intake and high frequency), and revealed a significant increase in assignment to the ‘risky’ drinking cluster in Veterans with a history of blast exposure, where the ‘risky’ cluster was characterized by a higher level of combat exposure and numbers of mTBIs with loss of consciousness (LOC). Together, these results suggest disparate trauma effects of single vs. repetitive blast exposure, where only repetitive blast mTBI is characterized by increased risky drinking self-report in Veterans and consumption front-loading in mice, and help to clarify the seemingly disparate findings in the literature regarding how mTBI may give rise to increased substance use/misuse.

## MATERIALS AND METHODS

### Animals and mouse model of blast overpressure

Male C57Bl/6 mice (Jackson Laboratory) aged 3–4 months (weight 25-33 g; mean 28.5 ± 0.17 g) were used. All animal experiments were carried out in accordance with Association for Assessment and Accreditation of Laboratory Animal Care guidelines and were approved by the VA Puget Sound Institutional Animal Care and Use Committees. The shock tube (Baker Engineering and Risk Consultants, San Antonio, TX) was designed to generate blast overpressures that mimic open field high explosive detonations encountered by military Servicemembers in combat, and the design and modeling characteristics have been described in detail elsewhere [18, 32, 33, 35]. Briefly, mice were anesthetized with isoflurane (induced at 5% and maintained at 2-3%), secured against a gurney, and placed into the shock tube oriented perpendicular to the oncoming blast wave (ventral body surface toward blast). Sham (control) animals received anesthesia only for a duration matched to blast animals. Repeated blast/sham exposures occurred successively over the course of three days (one per day). The blast overpressure (BOP) peak intensity (psi), initial pulse duration (ms), and impulse (psi▪ms) used were in keeping with mild blast TBI (19.9 psi +/-0.14 psi) [38, 39]. Under these experimental conditions, the overall survival rate exceeded 95%, with blast-exposed mice appearing comparable to sham-exposed mice by inspection 2-4 hours-post blast exposure as previously reported [18, 32-35]. All behavioral tests were conducted starting at one-month post sham/blast exposure.

### Low-dose ethanol-induced locomotor stimulation

Ethanol-induced locomotor stimulation was investigated over the course of three days. On day one, each animal was pre-exposed/habituated to the behavioral arena (clean rat cage) for 15 min. On the second day, animals were injected with saline (1.0 ml/kg w/v, i.p.) and were again allowed to explore the behavioral arena for 15 min. Finally, on day three, animals were injected with ethanol (2.0 g/kg, i.p.) and allowed to explore the behavioral area for 15 min. Activity was recorded from above and analyzed using Anymaze (Wood Dale, IL). Ethanol-induced locomotor stimulation was expressed as percent increase in distance traveled (ethanol/saline).

### High-dose ethanol-induced sedation, tolerance, and metabolism

Ethanol-induced sedation and tolerance were investigated using the loss of righting reflex (LORR) paradigm. Each animal was injected with ethanol (4 g/kg, i.p.) and upon sedation (1-3 minutes later) was placed on its back in a V-shaped trough (any animal that did not lose LORR within 5 min was excluded from the study (n=2)). Animals were then observed undisturbed for up to 5 hours. When the animal was able to right itself twice within 30 seconds (i.e., flipped from back to stomach in the V-shaped trough), total sedation time was recorded (LORR duration). The identical procedure was repeated 24 hours later to assess ethanol tolerance. Blood was collected from the submandibular vein at 10 min and 4 hours following injection to determine blood ethanol concentrations. Blood ethanol concentrations were determined using Bioassay Systems (Hayward, CA) EnzyChrome Ethanol Assay kit as per manufacturer instructions.

### Intermittent two-bottle choice (I2BC)

Starting 4 days prior to the commencement of the I2BC procedure, mice were singly housed, and their usual water bottle removed and replaced with two bottles filled each with water. The bottles were 50 mL centrifuge tubes with a #6.5 neoprene rubber stopper and 2.5” ball sipper tube (Antrin, Bellmore, NY). Bottles were weighed immediately prior to placement in cage and then again at 24-hour timepoints (at approximately 5:30 pm) throughout the duration of the study. Each time the bottles were weighed, their position in the cage was reversed to prevent any side bias. One night each week (Wednesday), bottles were also weighed at 8pm (2 hours into the dark cycle) to assess ‘binge’ like drinking patterns. Following 4 days of baseline exposure to the water bottles, animals had access to one water bottle and one ethanol bottle every Monday, Wednesday, and Friday, and access to only water every Tuesday, Thursday, Saturday, and Sunday. During the first week of ethanol exposure, the ethanol dose was slowly increased (3% on Monday, 6% on Wednesday, and 9% on Friday). For the remaining duration of the study (21 more days), the 20% ethanol dose was used. Body weight was recorded for each animal on Sundays and used to express ethanol intake in g/kg.

### Human subjects

These studies were approved by the VA Puget Sound Health Care System Human Subjects Committee and conformed to institutional regulatory guidelines and principles of human subject protection in the Declaration of Helsinki. All participants provided written informed consent prior to any study procedures. The current study used the self-reports of 105 Veterans with a history of blast exposure with acute symptoms and 34 deployed controls with no lifetime history of TBI of any severity (Supplementary Table 1). As previously reported [33], blast exposure(s) were reported as mild in nature and ranging from 1 (10%) to 50 or greater (15%) blast exposures. All participants in this report were male. Both males and females were eligible for enrollment and study inclusion. However, in this study we currently have no available data from blast-mTBI women because most mTBI Veteran participants were from combat military occupational specialties. Inclusion criteria for the blast group were as follows: documented hazardous duty in Iraq and/or Afghanistan with the U.S. Armed Forces and at least one blast exposure with acute symptoms (e.g., nausea, ringing in ears, blurry vision, hearing loss, unsteady on feet, eyes sensitive to light, headache, alteration of consciousness). The VA/Department of Defense/American Congress of Rehabilitation Medicine criteria was used to establish mTBI (for the blast group), with 97% of Veterans within this group meeting criteria for at least one mTBI. Exclusion criteria were as follows: moderate-severe TBI, seizure disorder, insulin-dependent diabetes, current diagnosis of alcohol or other substance abuse or dependence (excluding nicotine), schizophrenia or other psychotic disorders, bipolar disorder, dementia and taking medications likely to affect cognition performance. Inclusion and exclusion criteria for deployed control Veterans were identical with the exception that controls had no lifetime history of TBI of any severity. Exclusionary psychiatric disorders were ruled out by SCID-IV interview [40]. Self-report alcohol use was examined using the Alcohol Use Disorders Identification Test-Consumption Questions (AUDIT-C) [37].

### Unsupervised machine learning (cluster analysis)

Individual AUDIT-C question responses were used as features in an unsupervised machine learning model. Using Python and common scientific computing libraries (e.g., scipy, pandas, sk-learn), hierarchical clustering algorithm (Ward method) and k-means (Euclid) were used to assist in optimal cluster number selection. Silhouette, Davies Bouldin, and Calinski Harabasz scores were computed to assess k-means cluster assignments. Cluster stability was assessed using the scores for homogeneity, completeness, and mutual information criterion and a bootstrap approach with repeated random assignment of initial cluster centroids. K=3 clusters was chosen based on above evaluation metrics. Finally, cluster assignment was compared between deployed controls and Veterans with a history of blast exposure and potential effects of combat and blast exposure on cluster assignment was examined.

### Data analysis

As appropriate, data were analyzed using: (*i*) two-tailed Student’s t-tests; (*ii*) one-way or two-way (between/within subjects design) repeated measures analysis of variance (RM ANOVA), followed by Newman-Keuls Multiple Comparison Tests or Bonferroni post-hoc tests, respectively; (*iii*) Chi^2^ tests were conducted for categorical data (e.g., AUDIT-C). Reported *p* values denote two-tailed probabilities of p≤ 0.05 and non-significance (n.s.) indicates *p*> 0.05. Statistical analyses were conducted using Graph Pad Prism 4.0 (GraphPad Software, Inc., La Jolla, CA) and Python.

## RESULTS

### Ethanol-induced locomotor stimulation is prolonged following repetitive blast exposure

Depending on dose, ethanol can have stimulating and/or sedative effects, and can be assessed in mice via ethanol-induced locomotion and loss of righting reflex paradigms, respectively. To investigate whether blast exposure modulates the stimulating effects of ethanol, we first examined the ability of low-dose ethanol (2 g/kg) to increase locomotion (over saline) in C57BL/6 male mice one month after they received either one (1x) or three (3x, one per day) blast exposures. Ethanol stimulation was expressed as a percent change in distance traveled (ethanol/saline) and was examined in five-minute bins (15 min total). There were no statistically significant differences between 1x sham (n=8) and 3x sham-treated (n=9) mice (two-way RM ANOVA: interaction effect F[2,30]=0.554, p>0.05), thus, 1x and 3x sham animals were pooled together for subsequent analyses related to ethanol stimulation.

Figure 1a shows a significant effect of time post-ethanol injection (two-way RM ANOVA: main effect of time F[2,68]=19.54, p=0.0001, Bonferroni Multiple Comparison Test post hoc: n=8-17) and while post hoc analysis revealed no difference in locomotor stimulation at 5 min post ethanol injection, a significantly prolonged ethanol effect in 3x but not 1x animals was apparent when examined at 15 min post ethanol injection (Figure 1a). We next examined raw distance traveled during the entire 15 min tests across the three days of behavioral testing (habituation/baseline, saline, and ethanol) and found a significant effect across groups (two-way RM ANOVA: interaction effect F[4,68]=3.359, p=0.014, Bonferroni Multiple Comparison Test post hoc: n=8-17) (Figure 1b). Post hoc analyses demonstrated no differences during the initial habituation/baseline phase (base) but did reveal a significant increase in distance traveled during the final ethanol phase (EtOH) in 3x but not 1x animals.

**Figure 1:**
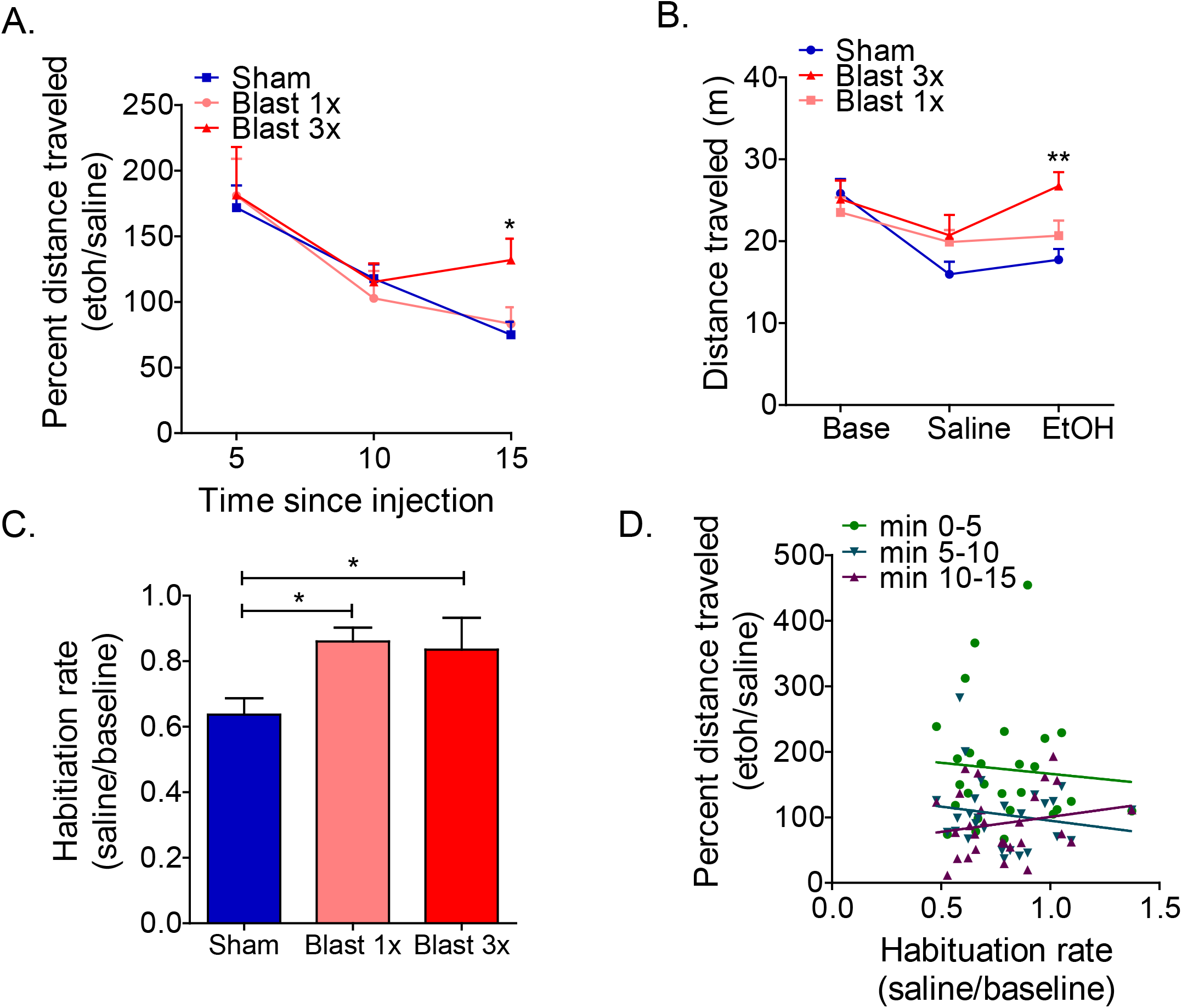
Repetitive blast exposure increases ethanol-induced locomotor stimulation. (a) Locomotor stimulating effects of ethanol as expressed by percent distance traveled. Two-way RM ANOVA *post hoc* Bonferroni Multiple Comparison Test. (b) Distance traveled at baseline and following saline or ethanol administration. Two-way RM ANOVA *post hoc* Bonferroni Multiple Comparison Test. (c) Locomotor habituation. One-way ANOVA *post hoc* Newman-Keuls Comparison Test. Correlation between ethanol locomotor stimulation and locomotor habituation rate. Spearman correlation. \**p* ≤ 0.05 \*\**p* ≤ 0.001: sham vs. blast. Values represent mean ± SEM.

Differences in locomotor habituation can potentially confound ethanol investigations and are especially relevant in paradigms where locomotion is repeatedly measured over a number of days, as is the case here. Indeed, when we computed a habituation rate (saline/base), we found a significant difference in both 1x and 3x blast exposed mice (one-way ANOVA: F[2,36]=4.946, p=0.013, Newman-Keuls Comparison Test post hoc: n=8-17) (Figure 1c). Critically, we did not find any significant correlation between habituation rate (saline/base) and ethanol stimulation (ethanol/saline) at any of the 5-min bins (min 0-5: Spearman ρ = 0.153, n.s, n=27; min 5-10: Spearman ρ = 0.749, n.s, n=27; min 10-15: Spearman ρ = 0.945, n.s, n=27), suggesting that our blast effects on ethanol stimulation were not mediated by potentially unrelated differences in locomotor habituation rates. Together these results demonstrate that repetitive blast exposure increases the duration of the stimulatory effects of ethanol, which is not related to blast-induced deficits in habituation.

### Blast exposure increases ethanol-induced sedation without affecting tolerance or metabolism

To investigate whether blast exposure modulates the sedative effects of ethanol, we examined the ability of high-dose ethanol (4 g/kg) to cause loss of righting reflex (LORR) on two consecutive days in C57BL/6 male mice one month after they received either one (1x) or three (3x, one per day) blast exposures. There were no statistically significant differences between 1x sham (n=8) and 3x sham-treated (n=9) mice (LORR day 1: Student’s unpaired t-test, t[16]=0.463, *p*>0.05; LORR day 2: Student’s unpaired t-test, t[16]=0.056, *p*>0.05), thus, 1x and 3x sham animals were pooled together for subsequent analyses related to ethanol sedation.

In accordance with previous results [25], a significant increase in LORR duration on the first day of testing was found in both 1x and 3x blast exposed mice (one-way ANOVA: F[2,42]=9.854, p=0.0003, Newman-Keuls Multiple Comparison Test post hoc: n=13-17) (Figure 2a). A similar increase was found on the second day of repeat testing (one-way ANOVA: F[2,42]=7.413, p=0.001, Newman-Keuls Multiple Comparison Test post hoc: n=13-17) (Figure 2b). We next assessed ethanol tolerance by examining LORR change (day 2 / day 1). Figure 2c shows significant tolerance to repeated ethanol injections for all groups (one-sample t-test vs. a theoretical mean of 1.0 (no tolerance): n=13-17; sham: t[16]=3.788, p=0.001, blast 1x: t[12]=3.709, p=0.003, blast 3x: t[12]=3.66, p=0.003), and no significant difference in tolerance rate across groups (one-way ANOVA: F[2,42]=1.615, p=0.211, n=13-17).

**Figure 2:**
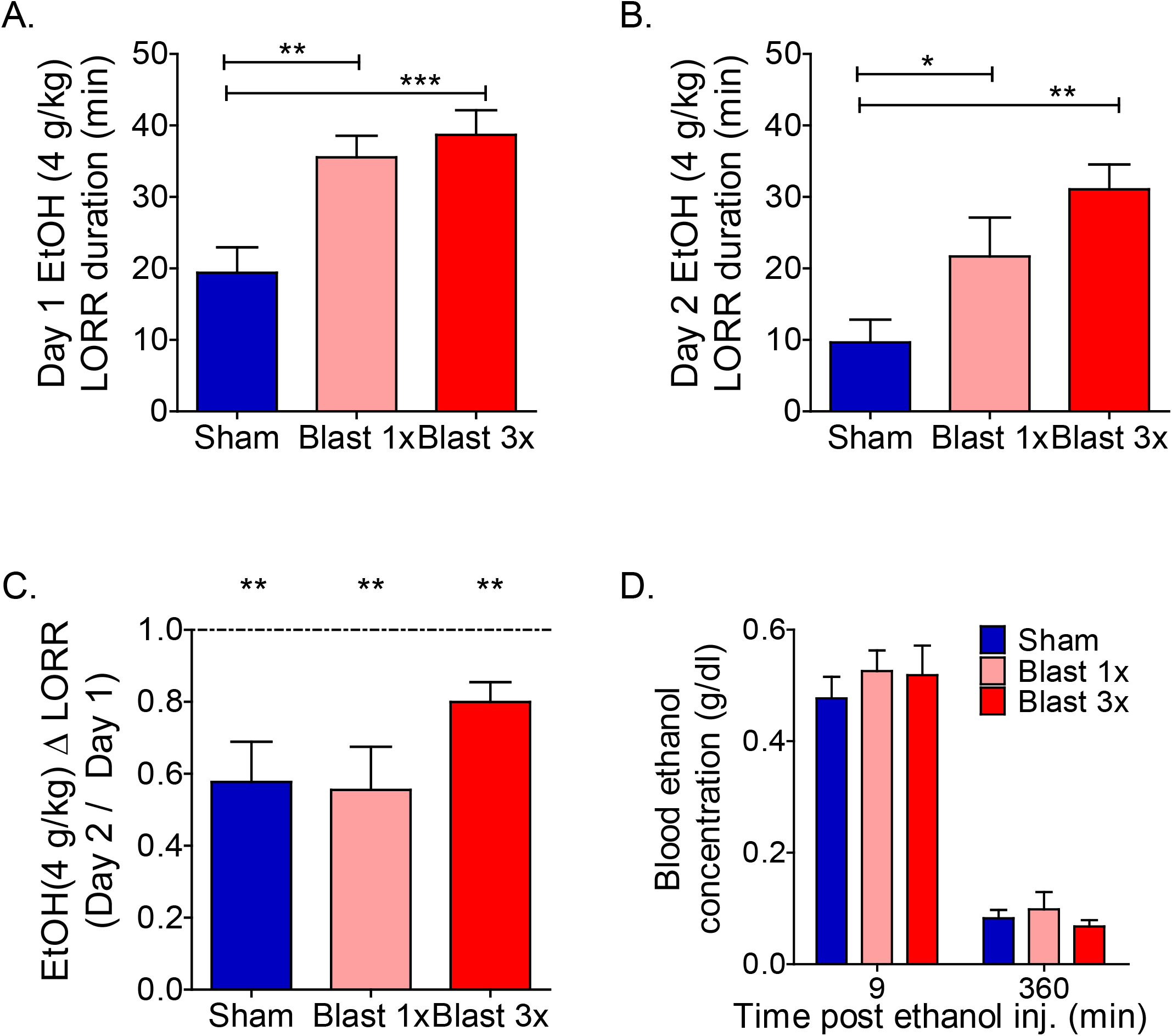
Blast exposure increases ethanol-induced loss of righting reflex. (a,b) Ethanol-induced LORR duration on day 1 (a) and day 2 (b) of ethanol administration. One-way ANOVA *post hoc* Newman-Keuls Comparison Test. (c) Tolerance to ethanol-induced LORR. One-way ANOVA *post hoc* Newman-Keuls Comparison Test. (d) Blood ethanol concentration 9 minutes and 4 hours after ethanol administration. Two-way RM ANOVA *post hoc* Bonferroni Multiple Comparison Test. \**p* ≤ 0.05, \*\**p* ≤ 0.001, and \*\*\**p* ≤ 0.0001. Error bars are mean +/-SEM.

Finally, we examined potential blast-induced changes to ethanol metabolism by measuring blood ethanol concentrations (BEC) at 10 min and 4 hours post injection. There were no statistically significant differences between 1x sham (n=4) and 3x sham-treated (n=5) mice (10 min: Student’s unpaired t-test, t[7]=1.667, *p*>0.05; 4 hour: Student’s unpaired t-test, t[7]=1.098, *p*>0.05), thus, 1x and 3x sham animals were pooled together for subsequent analyses related to ethanol metabolism. In accordance with previous results [25], no significant differences were found in ethanol metabolism at either time point in 1x or 3x blast exposed mice (two-way RM ANOVA: interaction effect F[2,15]=0.427, p>0.05, n=4-8) (Figure 2d).

### Repetitive, but not single, blast exposure decreases 24-hour ethanol intake but increases consumption ‘front-loading’

Previous results have suggested seemingly disparate trauma outcomes in rodents when ethanol self-administration is measured in short-access vs. long-access paradigms. To further investigate these phenomena, we conducted intermittent two-bottle choice testing with 20% ethanol in C57BL/6 male mice one month after they received either one (1x) or three (3x, one per day) blast exposures, where intake measurements were repeatedly taken at time points corresponding to short-(after approximately 2 hours of access) and long-access (after 24h of access). Consumption front-loading describes the tendency for some groups to consume significantly more in the acute phase of a self-administration paradigm after a period of forced abstinence and has implications for health-risk behaviors such as binge drinking.

Using this paradigm, repetitive, but not single, blast exposure caused a significant decrease in daily ethanol intake across the three weeks of testing (two-way RM ANOVA: main effect of group F[2,192]=4.417, p=0.023, n=8-10) (Figure 3a) and a significant decrease in average daily intake (one-way ANOVA: F[2,26]=4.632, p=0.02, Newman-Keuls Multiple Comparison Test post hoc: n=8-10) (Figure 3b). In accordance with these intake results, we found a similar pattern of effects when examining daily ethanol preference (ethanol intake/water intake) (two-way RM ANOVA: main effect of group F[2,192]=5.866, p=0.008, n=8-10) (Figure 3c) and average daily preference (one-way ANOVA: F[2,26]=6.038, p=0.007, Newman-Keuls Multiple Comparison Test post hoc: n=8-10) (Figure 3d) across the three weeks of testing.

**Figure 3:**
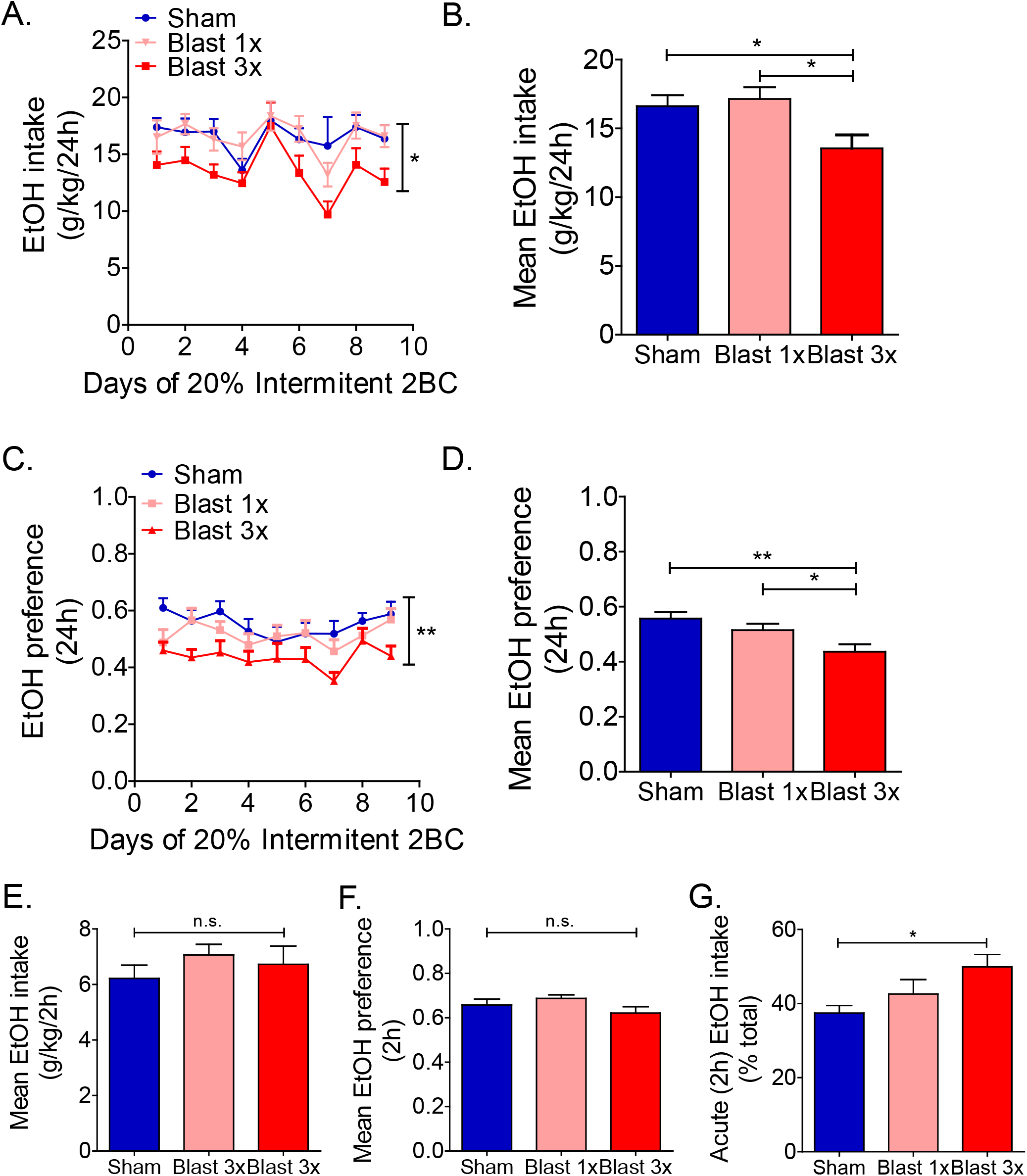
Blast exposure increases ethanol consumption ‘front-loading’. (a,b) Daily intermittent two-bottle choice 20% ethanol intake. Two-way RM ANOVA *post hoc* Bonferroni Multiple Comparison Test (daily) and One-way ANOVA *post hoc* Newman-Keuls Comparison Test (average). (c,d) Daily intermittent two-bottle choice 20% ethanol preference. Two-way RM ANOVA *post hoc* Bonferroni Multiple Comparison Test (daily) and One-way ANOVA *post hoc* Newman-Keuls Comparison Test (average). (e) Ethanol consumption during initial 2 hours of access. One-way ANOVA *post hoc* Newman-Keuls Comparison Test. (f) Ethanol preference during initial 2 hours of access. One-way ANOVA *post hoc* Newman-Keuls Comparison Test. Ethanol ‘front-loading’ (percent total intake). One-way ANOVA *post hoc* Newman-Keuls Comparison Test. \**p* ≤ 0.05 \*\**p* ≤ 0.001: sham vs. blast. Values represent mean ± SEM.

Conversely, when we measured intake levels approximately 2.5 hours after re-access to ethanol (i.e., each Wednesday evening at 6pm, 2 hours into the dark cycle). We found no significant difference between sham and blast-exposed animals in average intake (one-way ANOVA: F[2,26]=0.744, p=0.485; n=8-10) (Figure 3e) or preference (one-way ANOVA: F[2,26]=1.714, p=0.201; n=8-10) (Figure 3f) measured after approximately 2.5 hours of access. To assess potential changes in consumption ‘front-loading’ (e.g., the tendency to consume higher amounts immediately following a period of forced abstinence), we computed the percent of ethanol consumed within the first 2.5 hours of exposure and found that repetitive, but not single, blast exposure caused a significant increase in ‘front-loading’ consumption (one-way ANOVA: F[2,26]=3.975, p=0.032, Newman-Keuls Multiple Comparison Test post hoc: n=8-10) (Figure 3g). Together, these results suggest that repetitive blast exposure modifies the pattern of voluntary ethanol intake resulting in increased consumption ‘front-loading’.

### Cluster analysis reveals a shift in drinking patterns in Veterans following repetitive blast exposure

A cohort of 105 OEFOIF//OND Veterans with previous history of blast exposure with acute symptoms (BE) and 34 OEF/OIF/OND Deployed Control (DC) Veterans with no lifetime history of TBI of any severity were studied. Supplemental Table 1 shows that the BE and DC groups were statistically comparable in terms of age at time of evaluation, education level, and race distribution (non-white/white ratio). Veterans in the BE group had an average of 19 ± 3 blast exposures (median = 7) where the average time from their last blast exposure to evaluation was 5.4 ± 0.3 years. Current alcohol use was assessed as previously shown [15] with the Alcohol Use Disorders Identification Test-Consumption Questions (AUDIT-C), a screening measure used to identify individuals who are at risk for problematic drinking [37]. Scores on this measure range from 0-12, with 0 indicating no alcohol use and higher scores indicating more risk of unhealthy alcohol use. Previously established study criteria required individuals meeting DSM-IV criteria for alcohol abuse or dependence to be excluded from study. Nonetheless, AUDIT-C scores were significantly higher in BE as compared to DC (Chi^2^ = 20.95, p=0.021) (Figure 4a).

**Figure 4:**
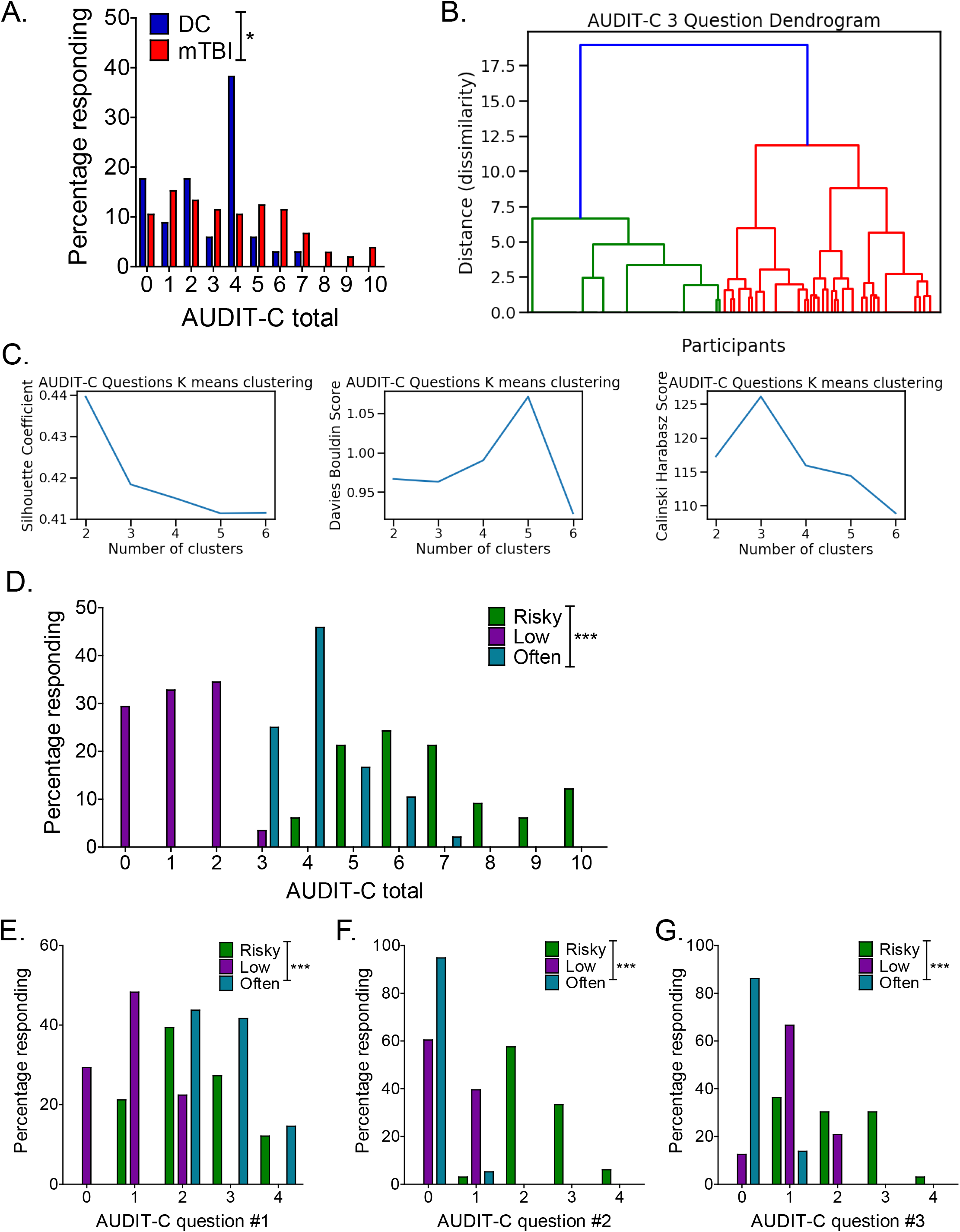
AUDIT-C cluster analysis. (a) Self-report AUDIT-C total scores in Veterans with/without a history of blast exposure with acute symptoms. Chi^2^. (b) Hierarchical clustering dendrogram visualization. (c) K-means cluster metrics highlight an optimal three cluster optimal. (d) AUDIT-C total scores across the three clusters. Chi^2^. (d) AUDIT-C total scores across the three clusters. Chi^2^. (e) AUDIT-C question 1 (i.e., drinking frequency) scores across the three clusters. Chi^2^. (f) AUDIT-C question 2 (i.e., drinking amount) scores across the three clusters. (g) AUDIT-C question 3 (i.e., binge-like drinking) scores across the three clusters. Chi^2^. \**p* ≤ 0.05 and \*\*\**p* ≤ 0.0001. Values represent mean ± SEM.

The AUDIT-C consists of three Likert-scale questions; while all three questions are related to intake patterns, each question probes a different drinking modality - the first question focuses on drinking frequency, the second on drinking quantity, and the third on “binge”-like drinking behavior. Here we used an unsupervised machine learning approach to examine potential sub-group differences across the AUDIT-C questions. Distance (dissimilarity) from a hierarchical clustering algorithm suggested an optimal cluster number of three (Figure 4b) and this was further supported by examining cluster metrics using a K-means clustering algorithm (Figure 4c). Finally, analysis of cluster stability also supported a three-cluster solution for this data set (Supplementary Table 2). Using k=3 with k-means clustering, Veterans were assigned to a single cluster based on their AUDIT-C responses. In support of this cluster assignment, total AUDIT-C scores were significantly different across cluster groups (Chi^2^ = 201.3, p=0.0001) (Figure 4d). Likewise, individual question AUDIT-C scores were also significantly different across cluster groups (question 1 (drinking frequency): Chi^2^ = 82.99, p=0.0001; question 2 (drinking quantity): Chi^2^ = 157.7, p=0.0001; question 3 (binge-like frequency): Chi^2^ = 123.4, p=0.001). Based on the distribution of scores within each cluster, we labeled the clusters as ‘low’ (n=58), ‘often’ (n=48), and ‘risky’ (n=33). Figure 4d-g show the outcomes of the cluster assignments: the ‘low’ drinking cluster is characterized by low responses across the three AUCID-C questions, the ‘often” drinking cluster is characterized by a high response on question 1 (i.e., the question related to drinking frequency) but low responses on questions 2 (i.e., quantity) and 3 (i.e., binge), and the ‘risky’ drinking cluster is characterized by high responses on questions 2 (i.e., the question related to drinking quantity) and 3 (i.e., the question related to binge-like consumption) and intermediate responses on question 1 (i.e., frequency).

Comparing cluster assignment between DC and BE Veterans, we found a significant change in cluster assignment (Chi^2^ = 6.326, p=0.042) towards an increased ‘risky’ group membership in the BE group (Figure 5a). To further understand potential drivers of cluster assignment, we examined whether there were significant differences in combat exposure and/or blast number across the clusters (focusing now only on BE Veterans). We found a significant increase in reported combat exposure (one-way ANOVA: F[2,103]=4.725, p=0.012, Newman-Keuls post hoc: n=29-43) (Figure 5b) within the ‘risky drinking cluster. Likewise, we found that the risky drinking cluster was also associated with significantly greater number of blast mTBIs resulting in loss of consciousness (one-way ANOVA: F[2,100]=5.21, p=0.007, Newman-Keuls Comparison Test post hoc: n=29-42) (Figure 5c), and greater number of blast mTBIs (one-way ANOVA: F[2,101]=3.895, p=0.023, Newman-Keuls Comparison Test post hoc: n=29-42) (Figure 5d). Together, these results suggest that repetitive blast mTBI increases alcohol intake and potentially risky drinking behaviors and are in correspondence with current results from our animal model of blast.

**Figure 5:**
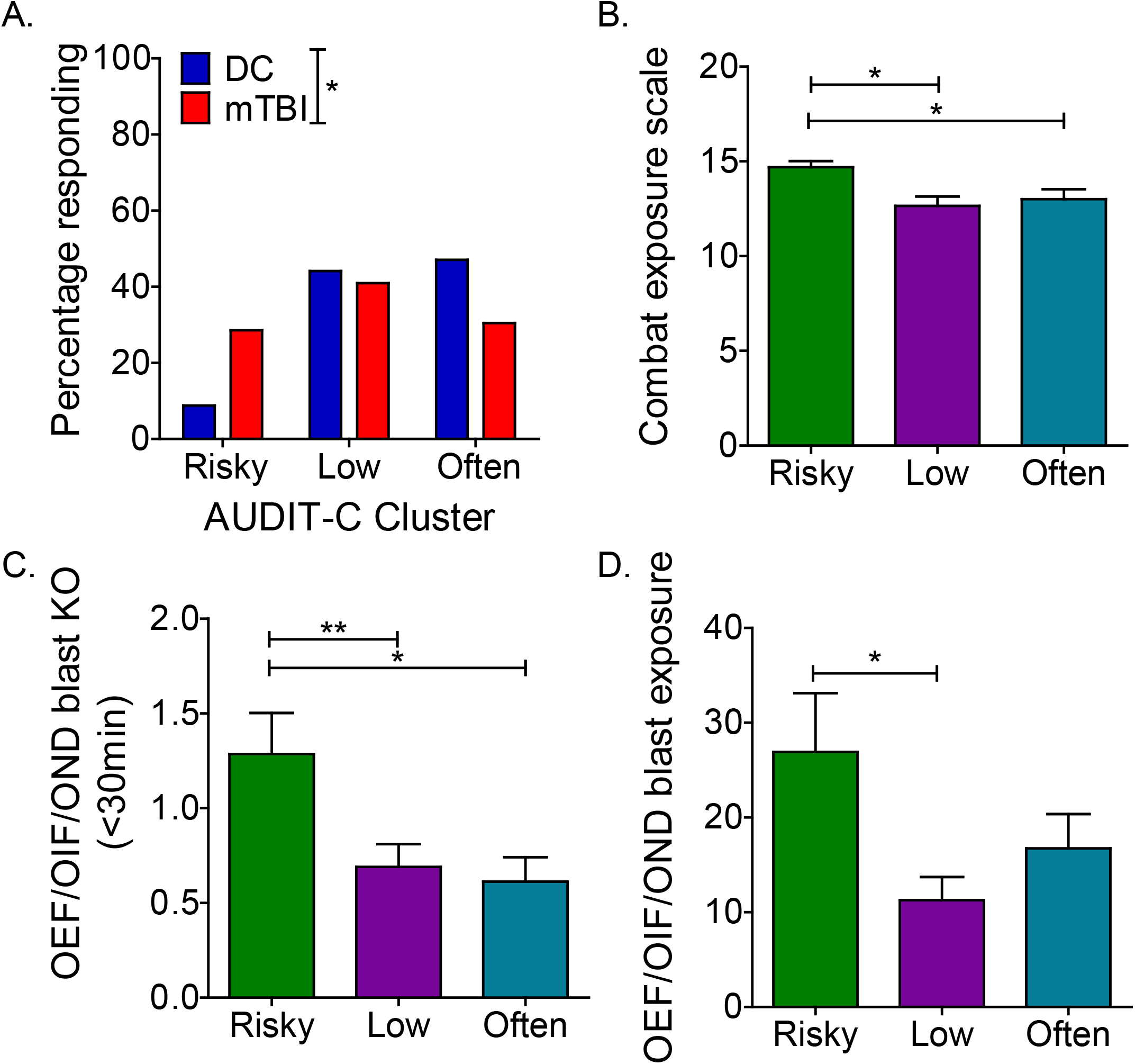
Blast mTBI results in shift to ‘risky’ drinking cluster. (a) AUDIT-C k-means cluster assignment in Veterans with/without a history of blast exposure with acute symptoms. Chi^2^. (b) Combat exposure by cluster assignment in Veterans with a history of blast exposure with acute symptoms. One-way ANOVA *post hoc* Newman-Keuls Comparison Test. (c) Blast mTBI with loss of consciousness (KO) count by cluster assignment in Veterans with a history of blast exposure with acute symptoms. One-way ANOVA *post hoc* Newman-Keuls Comparison Test. (d) Blast exposure count by cluster assignment in Veterans with a history of blast exposure with acute symptoms. One-way ANOVA *post hoc* Newman-Keuls Comparison Test. \**p* ≤ 0.05 and \*\**p* ≤ 0.01. Values represent mean ± SEM.

## DISCUSSION

Here we provide translational evidence demonstrating a blast-dose effect in relation to ethanol-locomotor stimulation and intake in mice, highlighting the importance of understanding blast mTBI history in OEF/OIF/OND Veterans that might be at heightened risk for health-risk behaviors related to substance misuse and/or abuse. Specifically, we found that while both single and repetitive blast exposure in mice increased ethanol sedation, only repetitive blast mTBI had significant effects on ethanol-induced locomotor stimulation and increased ethanol consumption ‘front-loading’. In corroboration, we found that Veterans with a history of blast with acute symptoms reported significantly more alcohol use as reported on the AUDIT-C. Using an unsupervised machine learning cluster analysis (k-means), a history of blast mTBI resulted in a significant shift to a ‘risky’ drinking cluster characterized by significantly higher blast mTBI numbers and blast mTBIs with loss of consciousness as compared to Veterans with a history of blast mTBI assigned to the ‘low’ and ‘often’ drinking clusters.

The armed conflicts of OEF/OIF/OND have resulted in an estimated mTBI rate of 10-25% with a comorbidity rate for PTSD of 50-75% [4-6]. In these conflicts, an estimated 75% of all TBIs reported by Servicemembers are a result of blast caused by detonation of high explosives [5, 9] and multiple deployments are common (2.77 million Servicemembers have served on 5.4 million deployments since 2011) [41], resulting in the increased potential for repetitive blast exposure. As such, blast exposure represents a major potential source of physical and psychological trauma, with implications for subsequent health-risk behaviors (e.g., sensation/novelty seeking, impulsivity, risk taking, irritability/aggression) and substance misuse/addiction. Indeed, both mTBI and PTSD can worsen pre-existing psychiatric disorders such as depression and increase and/or exacerbate substance misuse/addiction [22, 42] and other health risk behaviors [5, 10-18, 24], potentially compounding negative outcomes following injury and trauma. We previously reported increased PTSD and depression symptoms as well as alcohol use in OEF/OIF/OND Veterans with a history of blast mTBI as compared to deployed control Veterans with no lifetime history of TBI [11, 43], and more recently reported increased Veteran self-report of disinhibition and risk taking behaviors chronically following blast mTBI [18]. Importantly, therapeutic options following blast mTBI remain limited and are not universally effective.

Animal models support the notion of brain injury as a risk factor for adverse health-risk behaviors, including substance abuse and addiction [18, 24-30, 32, 44]. Using our established mouse model of repetitive blast exposure [33-35], we previously demonstrated increased novelty seeking in blast exposed mice [18] and more recently demonstrated acute stress responses and chronic PTSD-like outcomes following repetitive blast exposure [32]. Likewise, rats exposed to repetitive low-level blast developed a variety of anxiety, depression, and PTSD-related behavioral outcomes and potentiated responses to subsequent stressor exposure [30, 45, 46]. In relation to potential substance misuse and abuse, blast exposure with body shielding in rats increased voluntary ethanol intake during a short-access challenge session [27] and increased oxycodone seeking [26], and mild-to-moderate frontal cortical injury resulted in increased cocaine self-administration in rats [31] and increased ethanol sedation and decreased voluntary consumption (potentially due to increased ethanol sensitivity) in mice [25].

The underlying mechanisms by which repetitive blast drives subsequent health-risk behaviors and addiction risk remains unknown. Null results from the current study discount blast-induced changes to ethanol tolerance and metabolism as potential underlying mechanisms, which is in line with previous reports demonstrating that mTBI does not affect ethanol metabolism [25]. Conversely, we and others have demonstrated blast mTBI-induced changes to both tonic and phasic dopamine release patterns, as well as neuropathological/inflammatory changes within the mesolimbic system [18, 25, 31, 47-49]. We previously demonstrated a blast mTBI-induced increase in stimulated phasic dopamine release within the nucleus accumbens [18] and other reports demonstrate blast mTBI-induced neuroinflammation and tissue damage within the mesolimbic system [47-49]. Likewise, mild impact TBI models in rats and mice demonstrate alterations in mesolimbic dopamine receptors and related signal-transduction proteins [25, 31]. Damage to the mesolimbic system has been associated with deficits in executive function and emotional control, potentially leading to increased health-risk behaviors and addiction risk. Blast mTBI-induced changes to the structure and/or function of mesolimbic circuits thus pose a potential underlying mechanism related to blast-induced changes in ethanol and drug sensitivity and intake. While the adverse outcomes of trauma are thought to be mediated at least in part though maladaptive changes to the mesolimbic dopamine system [50-60], a causal role for mesolimbic dopamine dysfunction in blast mTBI-induced behavioral pathology has yet to be established and will be the focus of future investigations.

Our result of decreased daily ethanol intake one month following repetitive blast mTBI in mice might seem contrary to our clinical data suggesting increased ethanol intake self-report in Veterans with a history of blast mTBI. We interpret these findings to suggest that increased ethanol stimulation and sensitivity following repetitive blast exposure in mice acts to limit the overall amount of ethanol consumed (e.g., blast mTBI mice reach reward and/or intoxication more quickly than shams, thus limiting subsequent opportunities for consumption) [25]. In line with this idea, we found that ethanol consumption ‘front-loading’ was significantly increased in our blast mTBI mice, highlighting a potentially more “binge”-like intake pattern following repetitive blast exposure. Such ‘front-loading’ behavior is linked to ‘binge’-like consumption and has been previously reported in other animal models [61, 62] and observed in humans with increased vulnerability to developing problem drinking and alcohol use disorder [63]. These results from our animal model were corroborated by findings from our unsupervised cluster analysis of AUDIT-C self-report in Veterans with/without a history of blast mTBI. Specifically, frequency of assignment to the ‘risky’ drinking cluster was higher in Veterans reporting a history of blast mTBI with loss of consciousness (as compared to blast mTBI with only altered consciousness). Our cluster analysis findings are in line with a previous report demonstrating increased odds of frequent binge drinking in Veterans with a history of TBI with loss of consciousness as comparted to Veterans with no history of TBI or Veterans with history of TBI without loss of consciousness [23].

An AUDIT-C total score of 5 or higher in male Veterans is recommended as a positive screen for potential alcohol misuse and/or dependence (a score of 4 or higher is used for males in the general population), requiring follow-up with a health-care provider [37]. When examined for its predictive ability, a cut-off of 5 exhibits a high rate of specificity but lower sensitivity in its ability properly identify patients with substance misuse; indeed, this number was optimized to minimize the burden of false positive rates on VA providers [37]. Using our cluster based approach, the ‘frequent’ and ‘risky’ clusters share overlapping AUDIT-C scores of 4-7, raising the possibility that this approach might provide useful additional information to aid in the assessment of whether self-reported drinking behavior should be of concern for the medical provider. These results now require external validation in a larger sample without exclusion criteria related to substance abuse/dependence and/or in a population outside of the VA to determine potential merit of using such a cluster-based approach in a clinical care setting.

Together our results highlight that while a single mTBI can result in changes to the sedating properties of alcohol (without changes in tolerance or metabolism), repetitive mTBI results in prolonged ethanol-induced locomotor stimulation and ‘binge’-like consumption ‘front-loading’ in mice and a shift to the ‘risky’ drinking cluster in Veterans. Binge drinking specifically is associated with increased risk for negative consequences related to addiction, criminality, and chronic adverse health outcomes (e.g., obesity, liver damage). Over 400,000 OEF/OIF/OND Veterans have a history of blast exposure and mTBI (most often repetitive), highlighting the potential for significant costs related to blast mTBI-induced increases in risky drinking patterns and binging. While we are not able to draw causal inferences from our Veteran cohort, these data in combination with data from our animal model strongly support the notion of repetitive blast mTBI as a driver of health-risk behaviors such as risky drinking. Together, these results highlight the importance of understanding blast trauma history in OEF/OIF/OND Veterans who might also be at risk for substance misuse and/or abuse. Additional studies are warranted to further explore potential underlying mechanisms and treatment targets (e.g., the mesolimbic dopamine system) for those with a history of repetitive mTBI and higher likelihood of health-risk behaviors and substance misuse.

## Data Availability

Available upon request.

## Acknowledgments

This work was supported by a Department of Veteran Affairs (VA) Basic Laboratory Research and Development (BLR&D) Career Development Award 1IK2BX003258 (AGS), VA Clinical Sciences Research and Development (CSR&D) Career Development Award IK2CX001774 (RCH), a VA BLR&D Merit Review Award 5I01BX002311 (DGC), VA Rehabilitation Research and Development Service Merit Review Award #B77421 (ERP), University of Washington Friends of Alzheimer’s Research (DGC, ERP), UW Royalty Research Fund (DGC), the VA Northwest Mental Illness Research Education and Clinical Center (MAR, ERP, RCH), and the Burroughs Wellcome Fund Postdoctoral Enrichment Program grant #1019873 (BJ). We would like to thank Traci J Webber, Cindy Pekow, DVM, Kari Koszdin, DVM, and Monica Foley for considerable technical assistance and veterinary care.

## Disclaimer

The views expressed in this scientific presentation are those of the author(s) and do not reflect the official policy or position of the U.S. government or the Department of Veterans Affairs.

## Notes

### Competing Interest Statement

The authors have declared no competing interest.

### Author Declarations

VA Puget Sound IRB approved

