## Supplemental for "Repetitive but not single blast mild traumatic brain injury increases ethanol responsivity in mice and risky drinking behavior in combat Veterans"

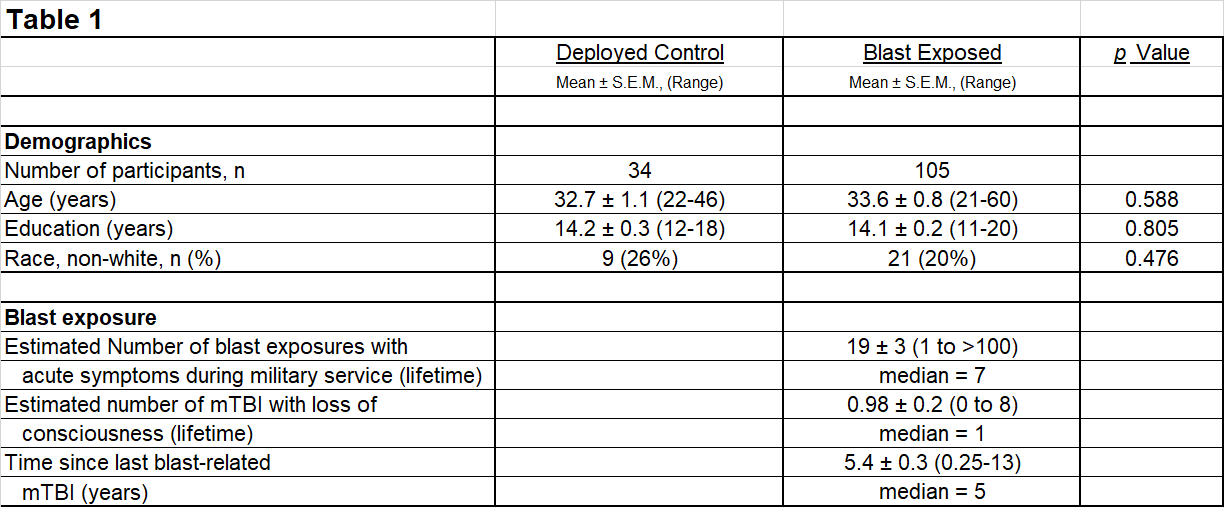
**Table 1: Study participant demographics and blast exposures.** Chi^2^.

**Table 2: K-means cluster stability analysis**.

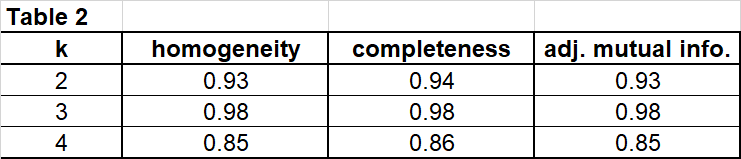
